# 24 People, one test: Boosting test efficiency using pooled serum antibody testing for SARS-CoV-2

**DOI:** 10.1101/2020.09.01.20186130

**Authors:** Stefan Nessler, Jonas Franz, Franziska van der Meer, Konstantina Kolotourou, Vivek Venkataramani, Chalid Hasan, Beatrix Pollok-Kopp, Andreas E. Zautner, Christine Stadelmann, Michael Weig, Stefan Pöhlmann, Markus Hoffmann, Joachim Riggert

**Affiliations:** Institute of Neuropathology, University Medical Center Göttingen, Göttingen, Germany; Institute of Pathology, University Medical Center Göttingen, Göttingen, Germany; Department of Transfusion Medicine, University Medical Center Göttingen, Göttingen, Germany; Institute of Medical Microbiology, University Medical Center Göttingen, Göttingen, Germany; Infection Biology Unit, German Primate Center - Leibniz Institute for Primate Research, Göttingen, Germany; Faculty of Biology and Psychology, Georg-August University Göttingen, Göttingen, Germany

## Abstract

**Background:** The global pandemic of COVID-19 (coronavirus disease 2019) is caused by the novel coronavirus SARS-CoV-2 (severe acute respiratory syndrome coronavirus 2), with different prevalence rates across countries and regions. Dynamic testing strategies are mandatory to establish efficient mitigation strategies against the disease; to be cost effective, they should adapt to regional prevalences. Seroprevalence surveys that detect individuals who have mounted an immune response against COVID-19 will help to determine the total number of infections within a community and improve the epidemiological calculations of attack and case fatality rates of the virus. They will also inform about the percentage of a population that might be immune against re-infections.

**Methods:** We developed a sensitive and specific cell-based assay to detect conformational SARS-CoV-2 spike (SARS-2-S) S1 antibodies in human serum, and have cross-evaluated this assay against two FDA-approved SARS-CoV-2 antibody assays. We performed pseudovirus neutralization assays to determine whether sera that were rated antibody-positive in our assay were able to specifically neutralize SARS-2-S. We pooled up to 24 sera and assessed the group testing performance of our cell-based assay. Group testing was further optimized by Monte Carlo like simulations and prospectively evaluated.

**Findings:** Highly significant correlations could be established between our cell-based assay and commercial antibody tests for SARS-CoV-2. SARS-2-S S1 antibody-positive sera neutralized SARS-2-S but not SARS-S, and were sensitively and specifically detected in pools of 24 samples. Monte Carlo like simulations demonstrated that a simple two-step pooling scheme with fixed pool sizes performed at least equally as well as Dorfman’s optimal testing across a wide range of antibody prevalences.

**Interpretation:** We demonstrate that a cell-based assay for SARS-2-S S1 antibodies qualifies for group testing of neutralizing anti-SARS-2-S antibodies. The assay can be combined with an easily implemented algorithm which greatly expands the screening capacity to detect anti-SARS-2-S antibodies across a wide range of antibody prevalences. It will thus improve population serological testing in many countries.

**Funding:** This work was supported by the Bundesministerium für Bildung und Forschung within the network project RAPID (risk assessment in pre-pandemic respiratory infectious diseases [grant number 01KI1723D, S.P.]).

## Introduction

The global pandemic of coronavirus disease 2019 (COVID-19) is caused by the severe acute respiratory syndrome coronavirus 2 (SARS-CoV-2)^1^. Appropriate mitigation strategies require the dynamic monitoring of disease propagation. RT-PCR screenings tend to underestimate the disease burden due to their inability to detect past infections. Furthermore, mildly affected or asymptomatic individuals are usually not tested. Seroprevalence surveys to assess the proportion of a population that has already developed anti-viral antibodies are less biased towards clinically severe cases. Their use contributed significantly to the delineation of COVID-19 transmission chains in Singapore^2^.

There are important global, national and regional differences in the prevalence of COVID-19, and diagnostic testing and screening strategies should be adapted to the geographic needs. The state of Lower Saxony, for example, is a low to medium prevalence area for COVID-19 in Germany (1683 cases per 10^6^ individuals), and individual testing to assess the seroprevalence of SARS-CoV-2 antibodies therefore might be unnecessarily time-consuming and expensive.

Group testing, which combines biological specimens from multiple individuals into one testing pool examined by a single test, has the potential to be both time- as well as resource-saving. The concept of screening sample pools was developed during World War II by Dorfman, who suggested that draftees be screened for syphilis antibodies in optimized group sizes for different prevalence rates (n=11 for 1%, n=8 for 2%)^3^. Back then, this algorithm was never implemented because the Wassermann test to detect syphilis antibodies was not sensitive enough when as few as eight samples were pooled. More recently, HIV antibodies were detected accurately in 10 sample mini-pools^4^, and group testing of blood donor sera is regularly performed in blood banks to search for common viral genomes by nucleic acid testing (NAT)^5^.

Here we present a cell-based antibody assay that specifically detects antibodies against the SARS-CoV-2 spike glycoprotein (SARS-2-S) S1 subunit. The assay is sensitive enough for group testing, and when combined with a simple, easily implemented algorithm, massively expands the screening capacity for the detection of SARS-CoV-2 antibodies in low and medium prevalence areas of the disease. Antibody-positive sera neutralize SARS-2-S in a pseudovirus neutralization assay. Thus, this approach can be of great value for determining the dynamics of anti-SARS-CoV-2 humoral immunity on a population level.

## Methods

### Patient characteristics

We first evaluated our cell-based assay with serum of individuals recruited by our Department of Transfusion Medicine as potential candidates to donate convalescent plasma. In this cohort, 54/61 individuals were tested positive for SARS-CoV-2 in combined nasal/ throat specimens, six had symptoms compatible with COVID-19 but had not undergone PCR testing, and one individual was PCR-negative. The female-to-male ratio was 1·77 and the median age was 42 years (IQR 31-54). The median time from clinical symptom onset to serum collection for SARS-CoV-2 antibody measurements was 63 days (IQR 56-81). At the time of antibody evaluations, 60/61 individuals were SARS-CoV-2 PCR-negative in combined nasal/ throat specimens and 1/61 test was inconclusive.

Furthermore, we collected 589 blood donor samples from March to June 2020 to evaluate the potential of the cell-based assay to reliably detect anti-SARS-CoV-2 antibodies in pools of 21-24 patients. In this cohort, the female-to-male ratio was 0-63 and the median age of this cohort was 48 (IQR 32-56).

Finally, we prospectively analyzed 30 NAT pools (28 × 24 sample pools; 1 × 18 sample pool; 1 × 14 sample pool; 704 individuals in total) obtained from the Department of Transfusion Medicine for the presence of anti-SARS-CoV-2 antibodies. In this cohort, the female-to-male ratio was 0-96 and the median age was 28 (IQR 23-44). Of note, some of the prospectively analyzed NAT pools contained sera from individuals donating convalescent serum.

Written consent was obtained from all individuals and the study was approved by the local ethics committee (14/8/20).

### Cell-based antibody measurements

HEK293 cells were stably transfected with either the pCMV3-2019-nCoV-Sl (HEK^spike S1^, Sino Biologicals VG40591-UT, China), pCMV3-HCoV-OC43-Spike-Flag (HEK^OC43^, Sino Biologicals, VG40607-CF) or the pCMV3-untagged (HEK^EV^, Sino Biologicals CV011) expression plasmids. HEK^spike S1^ cells with strong membrane staining by SARS-2-S S1 antibodies were enriched by cell sorting (BD FACSAria™ II) and cultivated further in DMEM, high glucose (Gibco™, Thermo Fisher Scientific, MA, USA) supplemented with 10% FCS (Gibco™), P/S (Gibco™) and 200 μg/ml hygromycin B (InvivoGen, CA, USA).

Individual human serum samples were pre-diluted 1 to 10 in DMEM growth medium and added to 20 000 HEK^spike S1^ or HEK^EV^ cells in 96 U wells, yielding a final serum dilution of 1 to 50 to measure IgG or IgG1 anti-SARS-2-S spike S1 antibodies. The plates were placed on ice and slowly shaken for 15 min. Cells were subsequently washed three times with washing buffer (PBS plus 2% FCS). Secondary fluorochrome-labelled antibodies were diluted in washing buffer and added for 15 min on ice. Cells were then washed three times, re-suspended in 150 μl washing buffer, and recorded on a BD LSR Fortessa™ cell analyzer. Results were analyzed using the FlowJo™ Software Version 7.6.1. The following secondary antibodies were used: F(ab’)2 goat anti-human IgG-APC (1:100, Jackson ImmunoResearch, PA, USA), anti-human IgG1 PE (1:100, clone HP6001, SouthernBiotech, AL, USA), anti-human IgG3 AF647 (1:100, clone HP6050, SouthernBiotech) and anti-human IgA APC (1:50, clone IS11-8E10, Miltenyi, Bergisch Gladbach, Germany). For the measurement of IgA or IgG3 anti-SARS-2-S S1 antibodies, human serum was diluted 1:25 or 1:5, respectively, to adjust for the reduced concentrations of these Ig isotypes in human serum.

For sample pooling, equal volumes of patient serum samples were mixed. Pooled samples were diluted 1 to 3 with HEK^spike S1^ or HEK^EV^ cells for 24 sample pools (final dilution of individual sera in this pool 1:72) or 1 to 12-5 for 4 sample pools (final dilution of individual sera 1:50).

The MFI ratios presented correspond to the median fluorescence intensity of HEK^spike S1^ divided by the median fluorescence intensity of HEK^EV^. Serum samples were rated antibody-positive if their MFI ratio exceeded the MFI ratio of controls + 5 SD.

### Abbott chemoluminescent microparticle immunoassay (CMIA) and Euroimmun ELISA

The Abbott SARS-CoV-2 IgG assay is a CMIA, which detects IgG antibodies against the SARS-CoV-2 nucleocapsid protein with excellent analytical performance^6^. The assay was run on the Abbott Architect instrument platform according to the manufacturer’s instructions. The SARS-CoV-2-IgG immunoassay by Euroimmun is a commercially available ELISA (Euroimmun; Lübeck, Germany #EI 2606-9601 G), which detects anti-SARS-CoV-2 spike S1 antibodies with high diagnostic accuracy^7^ and was used in previous seroprevalence studies^8^.

### NAT pools

NAT pools were generated automatically by a pipet robot (Tecan Genesis RSP150, Tecan, Switzerland) by pooling up to 24 individual samples of each 50 μl EDTA plasma. EDTA-Plasma was replaced by similar volumes of isotonic NaCl in NAT pools composed of less than 24 samples.

### SDS PAGE and Western Blot Analysis

Cells at 70-80% confluency were scraped into modified radioimmunoprecipitation assay (RIPA) lysis buffer (PBS pH=7·4, 0·5% sodium deoxycholate, 1% NP-40, supplemented with 1 mM orthovanadate, 1 mM PMSF and 1x complete Roche protease inhibitor cocktail). Total protein concentrations were measured by DC protein assay (Bio-Rad). Equal amounts of protein (15 μg) were heated at 95 °C for 5 min with SDS (4x buffer, Bio-Rad) or at 37 °C for 10 min with LDS Non-Reducing buffer (Thermo Fisher Scientific) for denaturating or non-denaturating conditions, respectively. The samples were run on Mini-PROTEAN TGX precast 4-15% gradient gels (Bio-Rad) at 90 V constant and transferred using the Bio-Rad nitrocellulose and Trans-Blot Turbo System. The membranes were blocked for 1 h in 10% non-fat milk powder in TBS-T and incubated overnight at 4 °C with primary antibodies. The following day, the membranes were washed with TBS-T and incubated with the secondary antibody. Proteins were detected with Western Lightning^®^ Plus ECL Enhanced Chemoluminescence Substrate (PerkinElmer) and the images were equally adjusted in Adobe Photoshop to enhance the visibility of the bands. The following primary and secondary antibodies were used: anti-SARS-CoV-2 spike protein S1 receptor binding domain (1:250, mouse monoclonal #1034515, R&D systems, MN, USA); anti-β-actin antibody AC15 (1:5000, Sigma-Aldrich, Merck, Darmstadt, Germany) and polyclonal rabbit anti-mouse Ig/HRP (1:1000, DAKO, Jena, Germany).

### Immunocytochemistry

70·000 HEK^spike S1^ or HEK^EV^ cells were grown overnight on poly-L-lysine-coated cover slips and subsequently incubated with COVID-19 serum diluted 1:200 in DMEM growth medium for 20 min at 4 °C. Cells were washed three times with PBS and then incubated for 15 min with a F(ab’)2 goat anti-human lgG-AF647 antibody (1:100, Jackson ImmunoResearch, PA, USA) at 4 °C. After three additional washing steps, the cells were fixed with 2% PFA at room temperature for 30 min and nuclei were counterstained with 4’, 6-diamidino-2-phenylindole (DAPI) at room temperature for 15 min. Cells were subsequently washed three times, cover slips were mounted on a glass microscope slide with DAKO fluorescence mounting medium and analyzed with an Olympus confocal microscope.

### Pseudotyped virus neutralization assay

Neutralizing activity in patient sera against SARS-2-S was assessed by employing a previously published protocol^9^. In brief, replication-deficient vesicular stomatitis virus (VSV) pseudotype particles (VSVpp,^10^) bearing either VSV glycoprotein (VSV-G), SARS-S or SARS-2-S were incubated with four-fold serial dilutions of heat-inactivated (30 min at 56 °C) patient serum (final serum dilutions: 1:25, 1:100, 1:400, 1:1600, 1:6400) or medium without serum (control) for 30 min at 37 °C, before the mixtures were inoculated on Vero 76 cells (kindly provided by Andrea Maisner, Philipps University Marburg) in 96-well plates. After an incubation period of 16 h at 37 °C, VSVpp cell entry was analyzed by measuring the activity of virus-encoded firefly luciferase in cell lysates using a Hidex Sense plate luminometer (Hidex) and the Beetle-Juice substrate (PJK GmbH). For normalization, VSVpp cell entry in the absence of serum was set as 100% and the relative VSVpp cell entry efficiency in the presence of the different serum dilutions was calculated. All sera were analyzed in technical triplicates.

### Statistical analysis

Normality tests (D’Agostino & Pearson omnibus normality test) were applied to the data sets. To compare two experimental groups, unpaired *t* tests were used for parametric data and Mann-Whitney U tests for non-parametric data. To compare three or more groups, one-way ANOVA with Tukey’s post test was performed for parametric data and the Kruskal-Wallis test with Dunn’s post test was applied for non-parametric data. Statistical significance was defined as p<005. Data are presented as mean ± SEM, if not otherwise stated.

### Monte Carlo like simulation

Monte Carlo like simulations of different prevalence rates were performed for an overall finite sample size of 1000 samples (10·000 realizations for each prevalence rate of 0·0025, 0·00385, 0·005, 0·0075, 0·01, 0·015, 0.02, 0·03, 0·04, 0·06, 0·08, 0·12, 0·16, 0·2, 0·32, 0·48 and 0·64 were tested for each pooling strategy). Pooling strategies were compared either to complete measurements or to the one step pooling strategy by Dorfman^3^.

## Results

### Detection of conformational human serum antibodies to SARS CoV-2 spike SI

To investigate serum antibody reactivity against conformationally preserved SARS CoV-2 spike S1 in COVID-19 patients, we transfected HEK293 cells with human SARS-2-S S1 glycoprotein (HEK^spike S1^) or with an empty vector (HEK^EV^). SARS-2-S S1 glycoprotein was expressed in HEK^spike S1^ transfected cells by immunoblot analysis (figure 1A) and detected in the cell membrane by immunocytochemistry or flow cytometry (figure 1B, C). The surface staining was highly reproducible at different serum dilutions, demonstrating that a cell-based assay with HEK^spike S1^ cells can be used to quantify antibody responses in COVID-19 patients (figure 1C). Representative histograms of serum samples with no, medium or high titer anti-SARS-2-S S1 antibodies are shown in figure 1D.

**Figure 1.**
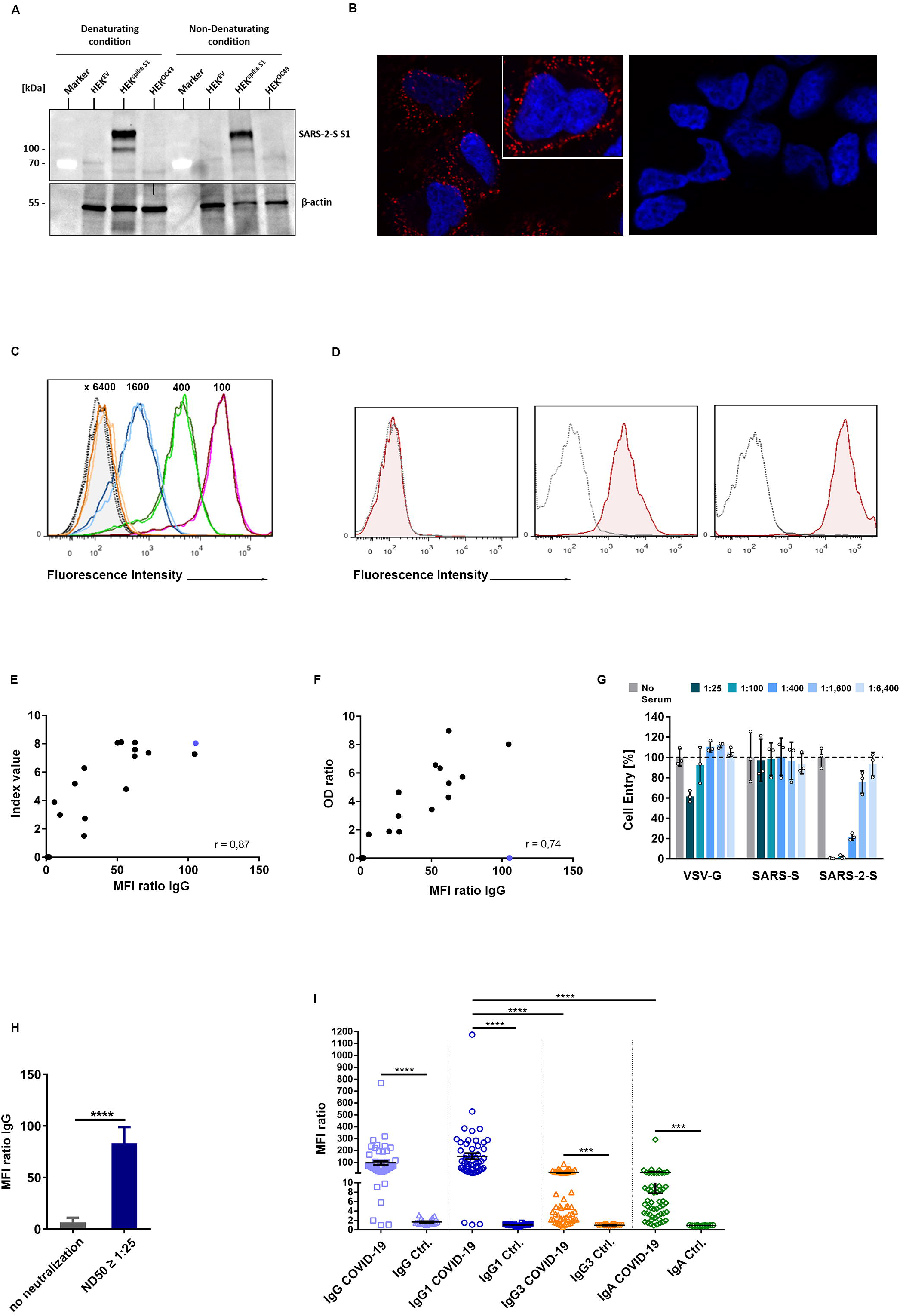
(A) Western blot analysis of SARS-2-S S1 glycoprotein expression in cell lysates from HEK^spike 1^ and HEK^EV^cells in reducing and non-reducing conditions. (B) Immunocytochemistry of HEK^spike 1^and HEK^EV^ cells by a COVID-19 serum. (C) Fluorescence intensity of HEK^spike 1^and HEK^EV^ cells analyzed by flow cytometry at different serum dilutions which are displayed on top of each curve. Duplicates are shown for each serum dilution. Dotted line represents serum antibody binding to HEK^EV^ cells. (D) Staining of HEK^spike 1^ (red line) and HEK^EV^ cells (grey line dotted) with sera of three representative patients at 1:50 dilution. Serum antibody binding was detected by an anti-human IgG 1 secondary antibody and quantified by flow cytometry. (E, F) Correlation of cell-based antibody measurements with two commercial FDA approved ELISAs (E, Abbott; p <0 0001; Spearman r=0-87; F, Euroimmun, p=0 0001; Spearman r=0-74). (G) Pseudovirus neutralization assay of the blue dotted serum, which was below the detection threshold of the Euroimmun assay but highly positive with both the Abbott CMIA and the cell-based antibody assay. (H) Column graphs depicting the MFI IgG ratios of sera with or without virus neutralizing activity (p <0 0001, Mann Whitney U test). (I) Comparative analysis of serum IgG, IgG1, lgG3 and IgA antibody titers to native spike S1 of COVID-19-positive patients and controls. Antibody binding to HEK^spik8 1^ and HEK^EV^ was determined by secondary fluorochrome-labelled antibodies targeting human IgG, IgG1, lgG3 or IgA and quantified by flow cytometry. Data are presented as median fluorescence intensity ratios (*** p <0 001, **** p <0 0001, Kruskal-Wallis test and Dunn’s post test).

We first cross-validated our cell-based assay with two commercial FDA-approved immunoassays, using a small serum (Abbott SARS-CoV-2 IgG) detects antibodies, which target the SARS-CoV-2 nucleocapsid protein. The CMIA by Abbott correlated strongly with the cell-based assay (figure 1E, p <0·0001, Spearman r=0·87) and all samples were equally rated either antibody-positive or -negative. Highly significant correlations could also be established for the Euroimmun assay (figure 1F, p=0·0001, Spearman r=0·74), but one of the 21 samples, strongly positive in our cell-based assay, was antibody-negative by Euroimmun standards. This particular serum (blue dot in figure 1E and F) had strong neutralizing capabilities in a pseudovirus-based neutralization assay with a neutralizing dose 50 (ND50) of 1:400 (see below and figure 1G), suggesting that our assay detects highly conformation-dependent antibodies.

### SARS-2-S S1 antibody-positive sera have neutralizing activity *in vitro*

We performed pseudovirus-based neutralization assays with SARS-2-S S1 antibody-positive and - negative sera. All sera lacking SARS-2-S S1 antibodies failed to inhibit the infection of Vero 76 cells with SARS-2-S pseudotyped VSV *in vitro* (figure 1H), suggesting that the presence of SARS-2-S S1 antibodies is biologically relevant. Two of 23 antibody-positive sera with low to medium antibody titers in our cell-based assay failed to neutralize SARS-2-S driven cell entry at a 1:25 dilution, and none of the SARS-2-S S1 antibody-positive or -negative sera neutralized SARS-S pseudotyped VSV *in vitro* (data not shown).

In summary, biologically relevant, virus-neutralizing antibodies against SARS-2-S S1 can be specifically detected by a cell-based HEK^spike S1^ assay with at least comparable sensitivity to the standard commercial immunoassays.

### IgG1 is the prominent Ig isotype of the anti-SARS-2-S S1 antibody response

We examined a large cohort of SARS-CoV-2 PCR+ individuals for the presence of anti-SARS-2-S S1 IgG antibodies at a median of 63 days after onset of COVID-19 symptoms. We found IgG antibodies to SARS-2-S S1 in 51/54 PCR+ patients. Further stratification of Ig isotypes demonstrated that IgG1 was the dominant isotype of the antibody response against SARS-2-S S1, with significantly higher MFI ratios compared to IgG3 or IgA (figure 1E, p <0·0001). Notably, none of the control samples collected before December 2019 harbored antibody reactivity against conformationally preserved SARS-2-S S1. Furthermore, the three sera with no antibody-reactivity in the cell-based SARS-2-S S1 assay, were also antibody-negative in the Abbott CMIA and Euroimmun ELISA.

### Cell-based assays detect SARS-S-2 S1 antibodies in pooled serum samples

We speculated that our highly sensitive cell-based assay might allow the pooling of multiple serum samples. To assess this, we first analyzed whether all SARS-2-S S1 antibody positive sera with low MFI ratios still exceeded the MFI ratios of healthy control pools by more than five standard deviations when mixed in equal volumes with 23 antibody-negative control sera. Indeed, all sera with established SARS-2-S S1 remained above the defined threshold (figure 2A). To test whether this approach might be used for population screening, we then pooled 21 to 24 sera of 589 blood donors collected between March and June 2020 to determine their MFI ratio in our cell-based assay. We identified 7/25 pools as being SARS-2-S S1 antibody-positive (figure 2B). Pool analysis was followed by the separate analysis of all blood donor sera in this cohort, and 9/589 sera were antibody-positive. The scatter plots presented in figures 2C and D illustrate that no antibody-positive serum was missed by this approach. For each pool, the MFI ratios of individual sera were measured and compared to the MFI ratios of the pooled measurements (size 21-24). The pooled MFI ratios correlated with the mean single MFI ratios when individual sera were positive (Figure 2C). Furthermore, the maximum MFI ratios of individual sera per pool were close to the MFI ratio of the pool (Figure 2D). The pooled MFI ratios did not correlate with the mean and maximum of the negative values when all sera were negative within a pool. In summary, cell-based antibody measurements qualify for group testing of SARS-2-S S1 antibodies.

**Figure 2.**
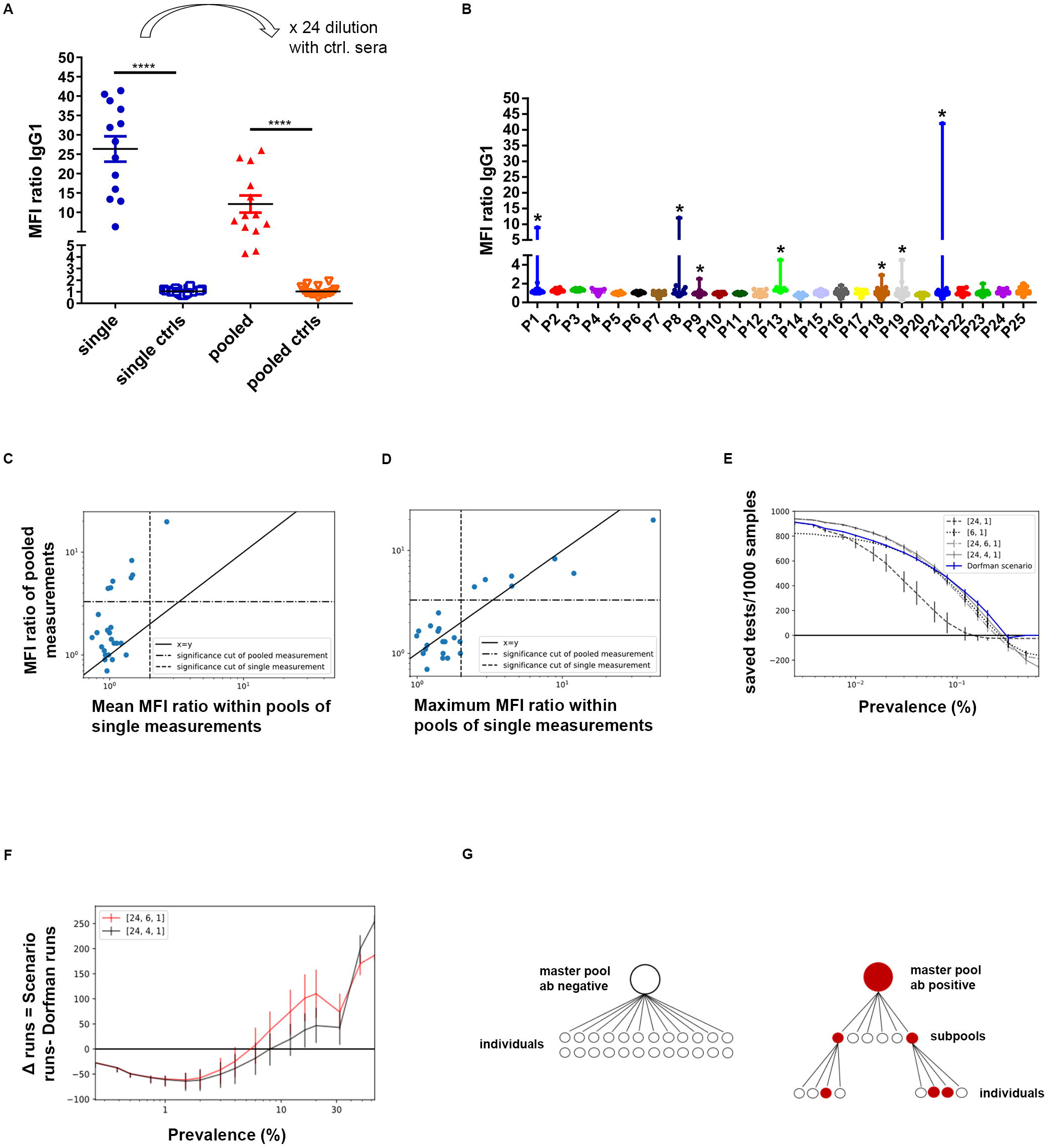
(A) Comparative analysis of individual and pooled antibody titers to native SARS-2-S S1. Antibody-positive sera with low MFI ratios were mixed with equal volumes of 23 antibody-negative control sera. Antibody binding to HEK^spike 1^ and HEK^EV^ of individual or pooled sera was determined by secondary anti-human IgG1 antibodies and quantified by flow cytometry (**** p <0 0001, one-way ANOVA and Tukey’s post test). (B) Whisker Plots with maximum values of 25 pools composed of 21-24 individual sera. Stars indicate, whether masterpools were determined antibody positive (> median FI ratio controls + 5 SD). (C, D) Scatter plots depicting mean (C) or maximum (D) MFI ratios of single measurements versus pooled MFI ratios. (E) Monte Carlo simulation of saved tests per 1000 samples assuming different prevalence rates. Error bars represent 95% confidence intervals according to 10000 sample distributions for each prevalence and proposed pooling strategy. (F) Saved tests compared to the Dorfman regime displayed by subtraction of required test numbers (Fixed regime minus Dorfman regime) for different prevalence rates. (G) Schematic of the testing algorithm.

We then optimized group testing by calculating pooling strategies with a mathematical search of all defectives in a binomial sample. Monte Carlo like simulation using different pooling schemes demonstrated that a simple, two-step pooling scheme with fixed pool sizes performed as well as Dorfman’s optimal testing across a wide range of antibody seroprevalences (figure 2E, F). We took advantage of NAT pools from blood donations, which we evaluated prospectively for the presence of anti-SARS-CoV-2 antibodies. Positive masterpools composed of up to 24 donor sera were subdivided, when positive, into six four-sample pools and subjected to further screening (figure 2G). As shown in figure 2F, at the given 1-5% prevalence of anti-SARS-CoV-2 antibodies in our region, subpools composed of 4 or 6 samples required comparable screening efforts. We found 11/30 pools to be antibody-positive, and each positive masterpool or four-sample subpool contained individual antibody-positive samples. In total, we performed 161 tests for the screening of 704 samples, and were thus able to demonstrate the rapid and sensitive screening for neutralizing antibodies in a large population-based cohort.

## Discussion

Most epidemiologists agree that wide-scale testing - also of asymptomatic individuals - is crucial for the safe re-opening of economies and societies. Seroprevalence surveys can inform policy makers about the percentage of people immune against re-infections at a given time point. These surveys might have to be repeated regularly if antibody titers drop over time, as suggested by Long and Seow et al.^11,12^. It is therefore essential that screening is readily accessible, and that the testing scheme can be efficiently adapted to regional antibody prevalences.

We decided to screen blood donors for the presence of anti-SARS-CoV-2 antibodies for a number of reasons. The well-established infrastructure of the Department of Transfusion Medicine, including standardized donor interviews as well as the generation of masterpools, which we received after NAT screening, was of great value. Also, blood donors are usually very committed and only seldom lost for follow-up evaluations; they thus provide a unique opportunity to follow antibody titers over time.

It is important to note, however, that blood donors differ from the general population in educational, social and health-related variables. Therefore, the prevalence rate of anti-SARS-CoV-2 antibodies obtained in this cohort might not entirely reflect the prevalence of anti-SARS-CoV-2 antibodies in the general population.

The highly significant correlation of the cell-based assay detecting SARS-2-S S1 antibodies with the Abbott CMIA detecting antibodies against SARS-2 nucleoproteins indicates that SARS-CoV-2 triggers a humoral immune response against at least two different epitopes comparably well. Cell-based assays are extraordinarily effective for detecting conformation-dependent antibodies, and serve as the standard method for measuring biologically relevant antibodies against surface antigens in patients with autoimmune disorders^13^. Accordingly, sera of COVID-19 patients that were antibody-positive in our SARS-2-S S1 cell-based assay demonstrated virus-neutralizing activity *in vitro*. The assessment of one serum sample of a COVID-19 PCR positive patient differed between the cell-based SARS-2-S S1 antibody assay, which was highly positive and the unremarkable, commercial SARS-2-S S1 antibody ELISA. This particular serum neutralized SARS-2-S-bearing pseudovirus particles very efficiently, suggesting that the patient’s anti-SARS-2-S S1 antibodies recognize conformationally sensitive epitopes that are lost when spotted onto ELISA plates.

We demonstrate here that individual anti-SARS-CoV-2 antibody-positive sera can be detected when mixed into pools of 24 samples. Of note, the pool size that can be assessed by this cell-based assay is more than twice as large as recently published pool sizes in group analyses of HIV antibodies^4^. Our method thus offers the unique possibility to perform group analyses for population screenings.

In the present study we chose a simple two-step approach with fixed pool sizes [24, 4, 1], which saves 64 (95 CI: -47, +83) tests compared to Dorfman’s optimal testing at the SARS-CoV-2 antibody prevalence of 1·5% in our area. This pooling scheme is at least comparably efficient to Dorfman’s optimal testing for antibody prevalences up to 10%. Thus, the testing scheme presented here is applicable across a wide range of antibody prevalences, and therefore may be useful in various epidemiological situations of the COVID-19 pandemic with diverse prevalence rates in different countries.

Sophisticated pooling schemes offering more efficient testing than the Dorfman scheme have been designed before^14,15^. Gosh and colleagues as well as Heidazardeh and Narayanan developed group testing schemes for RT-qPCR measurements of COVID-19 which use algorithms that also leverage soft information from the qPCR process to significantly improve Dorfman-style testing^16,17^. Thus it is clear that more complex testing algorithms will outcompete simple testing procedures. Notwithstanding, the high signal-to-noise ratio of cell-based anti-SARS-2-S S1 antibody measurements already allows quite efficient group testing even without the use of soft information.

In summary, we demonstrate a newly-developed cell-based assay that in combination with a simple algorithm offers efficient pool testing for neutralizing antibodies against SARS-CoV-2 across a wide range of antibody prevalences. In the present global health crisis, a population screening method with such time- and resource-saving potential could prove key for policy making in many countries.

## Data Availability

The raw data used in this study are available from the corresponding author upon reasonable request

## Contributors

SN designed the study. BPK and JR provided patient material and clinical data. SN, FvdM, KK, VV, CH, AEZ and MW performed the lab analyses. SP and MH provided the neutralization assays. JF performed the mathematical modelling. SN and CS wrote the first draft of the manuscript. All authors participated in the editing of the manuscript and approved the final version.

## Acknowledgements

The authors would like to thank all the participants of this study and express their gratitude for the excellent technical and administrative assistance of their institutes. We are indebted to Cynthia Bunker for language editing.

